# TyG-WC as a Clinical Tool for Hypertension Risk Stratification in Non-Diabetic Adults: A Large-Scale NHANES Analysis (2005-2016)

**DOI:** 10.1101/2025.07.05.25330939

**Authors:** Yuqiong Ou, Yuqiong Liu, Ping’an Chen, Chao Liu, Jing Li, Lanlan Ning, Lusi Chen, He-Jin LÜ, Zhen Liu

## Abstract

**Background/Objectives:** Hypertension risk stratification requires accessible metabolic tools. We evaluated three integrated indices that combine obesity and insulin resistance: TyG-BMI (triglyceride glucose-body mass index), TyG-WC (triglyceride glucose-waist circumference), and TyG-WHtR (triglyceride glucose-waist-to-height ratio). These indices were assessed for their ability to predict hypertension in non-diabetic adults.

**Methods:** Using nationally weighted data from 9, 593 NHANES participants (2005-2016), we applied survey-adjusted multivariable logistic regression and restricted cubic splines (RCS) to assess associations. Subgroup analyses further identified targets for precision interventions.

**Results:** TyG-WC showed superior prediction for hypertension, with a fully adjusted odds ratio (OR) of 4.09 (95% CI: 3.37–4.96). It exhibited a near-linear dose-response relationship (P for nonlinearity = 0.082), outperforming TyG-BMI (OR = 3.45, 95% CI: 2.83–4.22) and TyG-WHtR (OR=3.91, 95% CI: 3.20–4.78).Because of its simplicity—requiring only waist circumference, triglycerides, and glucose—and its linear relationship with hypertension risk, TyG-WC is suitable for use as a continuous clinical variable. Moreover, a TyG-WC cutoff of ≥600 offers optimal risk stratification, effectively replacing more complex metabolic syndrome criteria.Subgroup analyses revealed enhanced TyG-WC sensitivity in non-smokers (interaction P=0.012) and higher-educated individuals (P=0.037).

**Conclusions:** TyG-WC is a practical, cost-effective tool for hypertension screening in primary care. Its linear association allows direct integration into risk models. A value of ≥600 indicates the need for targeted intervention. Behavioral and sociodemographic factors further refine precision prevention strategies.

## Introduction

Hypertension is a major global public health challenge. It affects over 1.2 billion adults and contributes to severe cardiovascular and renal complications, which increase healthcare burdens[1]. Pharmacological and lifestyle interventions remain mainstays; however, their efficacy is limited by adherence issues and metabolic complexities[2]. This highlights the urgent need for simple and scalable risk prediction tools that can be deployed in primary care settings.

Insulin resistance (IR), a condition where cells respond poorly to insulin, is a central pathological link between metabolic dysfunction and hypertension[3].Hyperinsulinemia elevates blood pressure through sympathetic activation and endothelial impairment [4]. Dietary patterns, such as the Mediterranean diet rich in polyphenols, modulate the severity of insulin resistance [5, 6].However, traditional IR assessments, such as hyperinsulinemic-euglycemic clamps, are too costly and complex for routine use.

The triglyceride-glucose index (TyG) has emerged as a cost-effective IR surrogate [7], correlating with both metabolic syndrome and hypertension screening potential [8]. Critically, TyG-derived composites—TyG-BMI (general adiposity), TyG-WC (visceral fat), and TyG-WHtR (central obesity)—offer comprehensive insights into metabolic-lipid dysregulation [9, 10].For instance, TyG-WC reflects the combined impact of visceral adiposity and dyslipidemia that contribute to hypertension [11].

However, existing studies have notable limitations:

(1) Focus mainly on general populations, ignoring non-diabetic metabolic risk;
(2) Lack comparative analyses of TyG composites for precise screening;
(3) Insufficient use of national data for establishing generalizable thresholds.

To address these gaps, we use the NHANES database (2005-2016) to:

(1) systematically compare TyG-BMI, TyG-WC, and TyG-WHtR for predicting hypertension in non-diabetic adults;
(2) identify high-risk subgroups that may benefit from TyG-based interventions.

Our goal is to develop these indices into practical tools for healthcare systems with limited resources.

## Materials and methods

### Study design and data source

This study used cross-sectional data from the National Health and Nutrition Examination Survey (NHANES, 2005-2016). It aimed to explore the association between nutrition-metabolism-related indices (TyG-BMI, TyG-WC, TyG-WHtR) and hypertension risk. NHANES uses a complex, multistage stratified sampling design to collect demographic, lifestyle, dietary, clinical, and laboratory data from a nationally representative sample of the U.S. non-institutionalized population. To reflect the true distribution of the target population, the analysis incorporated NHANES-provided examination weights. It also adjusted for complex survey design effects using stratification (SDMVSTRA) and clustering variables (SDMVPSU). The study protocol adhered to ethical standards, and all participants provided written informed consent [12]. As a result, no additional informed consent or ethical review was required for this study. Data were obtained from the NHANES public database (available at https://www.cdc.gov/nchs/nhanes/).

### Study population and exclusion criteria

Starting with 33, 832 participants from NHANES 2005-2016, we applied the following exclusion steps (Fig 1):

**Fig 1.**
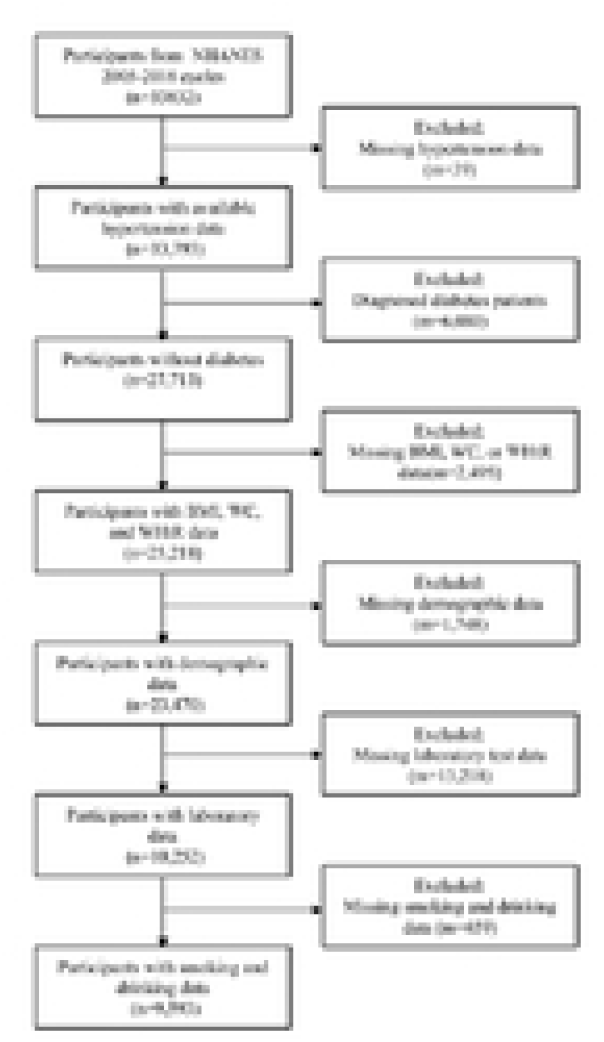
Flow chart of participant selection. NHANES, National Health and Nutrition Examination Survey. BMI, body mass index; WC, waist circumference; WHtR, waist to height ratio.

#### Exclusion of diabetic individuals

Based on the American Diabetes Association (ADA) criteria, participants have been excluded if they meet any of the following: self-reported physician-diagnosed diabetes, use of insulin or oral hypoglycemic agents, fasting blood glucose (FBG) ≥7.0 mmol/L, or HbA1c ≥6.5% (n=6, 080) [13].

#### Exclusion due to missing key variables

Participants with missing data on hypertension status (n=59), body mass index (BMI), waist circumference (WC), or waist-to-height ratio (WHtR) (n=2, 495) were excluded. Additionally, those missing demographic information (n=1, 748), laboratory parameters (n=13, 218), or smoking/drinking behaviors (n=659) were also excluded. The final analytical sample included 9, 593 non-diabetic adults aged between 20 and 80 years.

### Variable definitions and measurements

#### Primary exposure variables

TyG Index = ln[(TG [mmol/L] × 88.57) × (FBG [mmol/L] × 18)] / 2 [14].

This index combines fasting triglycerides (TG) and glucose to indicate dysregulated glucose-lipid metabolism.

Composite TyG indices include:

TyG-BMI = TyG × BMI (kg/m²), reflecting systemic obesity and metabolic dysregulation [15];
TyG-WC = TyG × WC (cm), highlighting visceral adiposity-driven metabolic risk [16];
TyG-WHtR = TyG × WHtR (waist circumference/height ratio), assessing central obesity and fat distribution [17].

#### Outcome variable (Hypertension)

The outcome variable (hypertension) was defined according to the 2017 ACC/AHA guidelines [18] by meeting any of the following criteria:

Systolic blood pressure (SBP) ≥140 mmHg or diastolic blood pressure (DBP) ≥90 mmHg; self-reported physician diagnosis; current use of antihypertensive medications.

#### Covariates

Based on previous studies [19–20], this research collected important covariates, including demographic, behavioral, and laboratory data.

Demographics: age, gender, race (Mexican American, Other Hispanic, Non-Hispanic White, Non-Hispanic Black, Other Races), and education (high school and below/college and above).

Nutritional and Metabolic Parameters: body mass index (BMI), waist circumference (WC), waist-to-height ratio (WHtR), high-density lipoprotein cholesterol (HDL-C), low-density lipoprotein cholesterol (LDL-C), fasting triglycerides (TG), fasting blood glucose (FBG), and insulin resistance index (INS).

Behavioral risk factors:

Smoking: Defined as smoking ≥100 cigarettes in one’s lifetime (yes/no). Drinking: Defined as consuming ≥12 alcoholic beverages per year (yes/no).

### Statistical analysis

#### Data description and weighting

Continuous variables were described as weighted mean ± standard error for normally distributed data or median (interquartile range), and categorical variables were reported as weighted percentages.

#### Group comparisons

To further investigate the association between triglyceride-glucose (TyG)-related indices and hypertension, we divided all patients into three groups. These groups were based on the quintiles of TyG-BMI, TyG-WC, and TyG-WHtR. The first quintile was defined as Q1, the second and third quintiles as Q2, and the fourth and fifth quintiles as Q3.Differences between groups were analyzed using the survey-weighted chi-square test for categorical variables or the design-adjusted Kruskal-Wallis test for non-normally distributed continuous variables [21].

#### Multivariable logistic regression

Three nested models were constructed: Model 1 (Crude) unadjusted; Model 2 (Adjusted model) adjusted for age and gender; and Model 3 (Fully Adjusted) further adjusted for race, education, smoking, and drinking. The results are presented as weighted odds ratios (ORs) with 95% confidence intervals (CIs).

#### Dose-response analysis

Restricted cubic splines (RCS, 4 knots) were incorporated into multivariable logistic regression models to evaluate nonlinearity associations between TyG indices and hypertension risk, with significance tested using likelihood ratio tests (P for nonlinearity <0.05).

#### Subgroup and interaction analyses

Stratified analyses were conducted based on age (<45/≥45 years), gender, race, education, smoking, and drinking to assess effect heterogeneity.

#### Software and significance

Analyses were performed using R version 4.2.1. A two-sided P value less than 0.05 was considered statistically significant.

## Results

### Metabolic characteristics and distribution of nutrition-related indicators in the study population

This study included 9, 593 non-diabetic adults (3, 391 in the hypertension group and 6, 202 in the non-hypertension group). Comparative analysis of baseline characteristics revealed significant associations between hypertension and components of metabolic syndrome. Key findings are summarized in Table 1:

**Table 1.** Baseline characteristics of study population by hypertension status.

| Characteristics | Total<br>(n=9593) | No-hypertension<br>(n=6202) | Hypertension<br>(n=3391) | P<br>value |
| --- | --- | --- | --- | --- |
| Age (years),<br>mean±sd | 44.98 ± 16.42 | 40.19 ± 14.64 | 55.05 ± 15.35 | <0.001 |
| Gender, n(%) |  |  |  |  |
| Male | 4687 (48.9) | 2965 (47.8) | 1722 (50.8) | 0.106 |
| Female | 4906 (51.1) | 3237 (52.2) | 1669 (49.2) |  |
| Race, n(%) |  |  |  |  |
| Mexican<br>American | 1502 (15.7) | 1136 (18.3) | 366 (10.8) | <0.001 |
| Other Hispanic | 966 (10.1) | 672 (10.8) | 294 ( 8.7) |  |
| Non-Hispanic<br>White | 4467 (46.6) | 2753 (44.4) | 1714 (50.5) |  |
| Non-Hispanic<br>Black | 1755 (18.3) | 986 (15.9) | 769 (22.7) |  |
| Other race | 903 ( 9.4) | 655 (10.6) | 248 ( 7.3) |  |
| Education, n(%) |  |  |  |  |
| High school and<br>below | 4373 (45.6) | 2638 (42.5) | 1735 (51.2) | <0.001 |
| College and<br>above | 5220 (54.4) | 3564 (57.5) | 1656 (48.8) |  |
| SBP (mmHg),<br>mean±sd | 120.16 ± 16.16 | 114.38 ± 10.61 | 132.30 ± 18.86 | <0.001 |
| DBP (mmHg),<br>mean±sd | 69.49 ± 11.90 | 67.64 ± 9.82 | 73.38 ± 14.63 | <0.001 |
| WT (kg),<br>mean±sd | 81.08 ± 20.43 | 78.77 ± 19.21 | 85.95 ± 22.00 | <0.001 |
| HT (cm),<br>mean±sd | 169.31 ± 9.92 | 169.48 ± 9.73 | 168.95 ± 10.31 | 0.027 |
| WC (cm),<br>mean±sd | 97.01 ± 15.63 | 94.29 ± 14.92 | 102.73 ± 15.54 | <0.001 |
| WHtR,<br>mean±sd | 0.57 ± 0.09 | 0.56 ± 0.09 | 0.61 ± 0.09 | <0.001 |
| BMI (kg/m<sup>2</sup>),<br>mean±sd | 28.19 ± 6.35 | 27.34 ± 5.96 | 29.99 ± 6.76 | <0.001 |
| HDL-C<br>(mmol/L),<br>mean±sd | 1.43 ± 0.42 | 1.44 ± 0.41 | 1.41 ± 0.43 | 0.016 |
| LDL-C<br>(mmol/L),<br>mean±sd | 2.99 ± 0.89 | 2.96 ± 0.87 | 3.05 ± 0.93 | 0.001 |
| TG (mmol/L),<br>mean±sd | 1.30 ± 0.73 | 1.22 ± 0.69 | 1.47 ± 0.79 | <0.001 |
| FBG (mmol/L),<br>mean±sd | 5.44 ± 0.54 | 5.34 ± 0.51 | 5.65 ± 0.55 | <0.001 |
| TyG, mean±sd | 8.49 ± 0.56 | 8.41 ± 0.55 | 8.66 ± 0.55 | <0.001 |
| TyG-WC, mean±sd | 826.82 ± 161.36 | 795.96 ± 153.70 | 891.70 ± 157.83 | <0.001 |
| TyG-BMI, mean±sd | 240.39 ± 60.69 | 230.87 ± 56.83 | 260.38 ± 63.62 | <0.001 |
| TyG-WHtR, mean±sd | 4.89 ± 0.94 | 4.70 ± 0.89 | 5.28 ± 0.92 | <0.001 |
| FINS (pmol/L), mean±sd | 66.83 ± 54.84 | 61.14 ± 49.01 | 78.79 ± 63.80 | <0.001 |
| Drinking, n(%) |  |  |  |  |
| Yes | 7098 (74.0) | 4673 (75.3) | 2425 (71.5) | 0.005 |
| No | 2495 (26.0) | 1529 (24.7) | 966 (28.5) |  |
| Smoking, n(%) |  |  |  |  |
| Yes | 4285 (44.7) | 2576 (41.5) | 1709 (50.4) | <0.001 |
| No | 5308 (55.3) | 3626 (58.5) | 1682 (49.6) |  |

n, number; SBP, systolic blood pressure; DBP, diastolic blood pressure; WT, weight; HT, height; WC, waist circumference; WHtR, waist-to-height ratio; BMI, body mass index; HDL-C, high-density lipoprotein cholesterol; LDL-C, low-density lipoprotein cholesterol; TG, triglyceride; FBG, fasting blood glucose; TyG, triglyceride glucose index; TyG-BMI, triglyceride glucose-body mass index; TyG-WC, triglyceride glucose-waist circumference; TyG-WHtR, triglyceride glucose-waist to height ratio; FINS, fasting insulin. P<0.05 was considered statistically significant.

#### Demographic Factors

The hypertension group had a markedly older mean age (P < 0.001) and a higher proportion of individuals with lower education levels (P < 0.001), emphasizing the role of health literacy in blood pressure management.

#### Central Obesity Indicators

Central obesity markers, including waist circumference (WC), waist-to-height ratio (WHtR), and body mass index (BMI), were significantly elevated in the hypertension group (all P < 0.001), indicating visceral adiposity as an independent risk factor for hypertension.

#### Glucose Metabolism Abnormalities

Fasting blood glucose (FBG) and TyG index levels were significantly higher in the hypertension group (P < 0.001), reflecting a strong link between insulin resistance (IR) and hypertension.

#### Lipid Profile Dysregulation

The hypertension group exhibited higher triglyceride (TG) levels (P < 0.001), lower HDL-C (P = 0.016), and slightly elevated LDL-C (P = 0.001), suggesting that lipid metabolism disorders may contribute to atherosclerotic processes.

#### Insulin Resistance Markers

Composite indices of insulin resistance (TyG-WC, TyG-BMI, TyG-WHtR) and fasting insulin (FINS) levels were significantly elevated in the hypertension group (all P < 0.001), supporting their utility as predictive biomarkers for hypertension.

#### Behavioral Risk Factors

Smoking prevalence was higher in the hypertension group (P < 0.001), while the prevalence of drinking was slightly lower (P = 0.005), which may reflect health awareness or reverse causality.

## Conclusion

Hypertension is closely associated with modifiable metabolic abnormalities and unhealthy lifestyle behaviors. These findings highlight the importance of early intervention and integrated health management strategies targeting visceral obesity, insulin resistance, and other risk factors.

### Differences among groups were compared according to TyG-related indicators

Tables 2-4, This study systematically evaluated the association between TyG-BMI, TyG-WC, and TyG-WHtR and the incidence of hypertension. The results showed that as these three composite metabolic indicators increased, both SBP and DBP rose significantly. For example, in the TyG-WC Q3 group, the median SBP was 122 mmHg and the median DBP was 72 mmHg. In contrast, the Q1 group had median SBP and DBP of 112 mmHg and 66 mmHg, respectively. These differences were statistically significant (P<0.001). This indicates that abdominal obesity, along with general obesity and metabolic disorders, is closely related to the incidence of hypertension.Furthermore, individuals in the Q3 group showed more severe insulin resistance and lipid abnormalities, including higher TyG index and fasting insulin, lower HDL-C, and elevated TG. These findings suggest that insulin resistance and lipid imbalance are key mechanisms driving hypertension development.Among individuals with elevated metabolic indicators, a higher proportion were male, Mexican American, and had lower education levels. This suggests an interaction between demographic characteristics and metabolic risk. Additionally, smoking rates were significantly higher in the TyG-WC and TyG-WHtR Q3 groups, potentially further increasing hypertension risk.

**Table 2.**
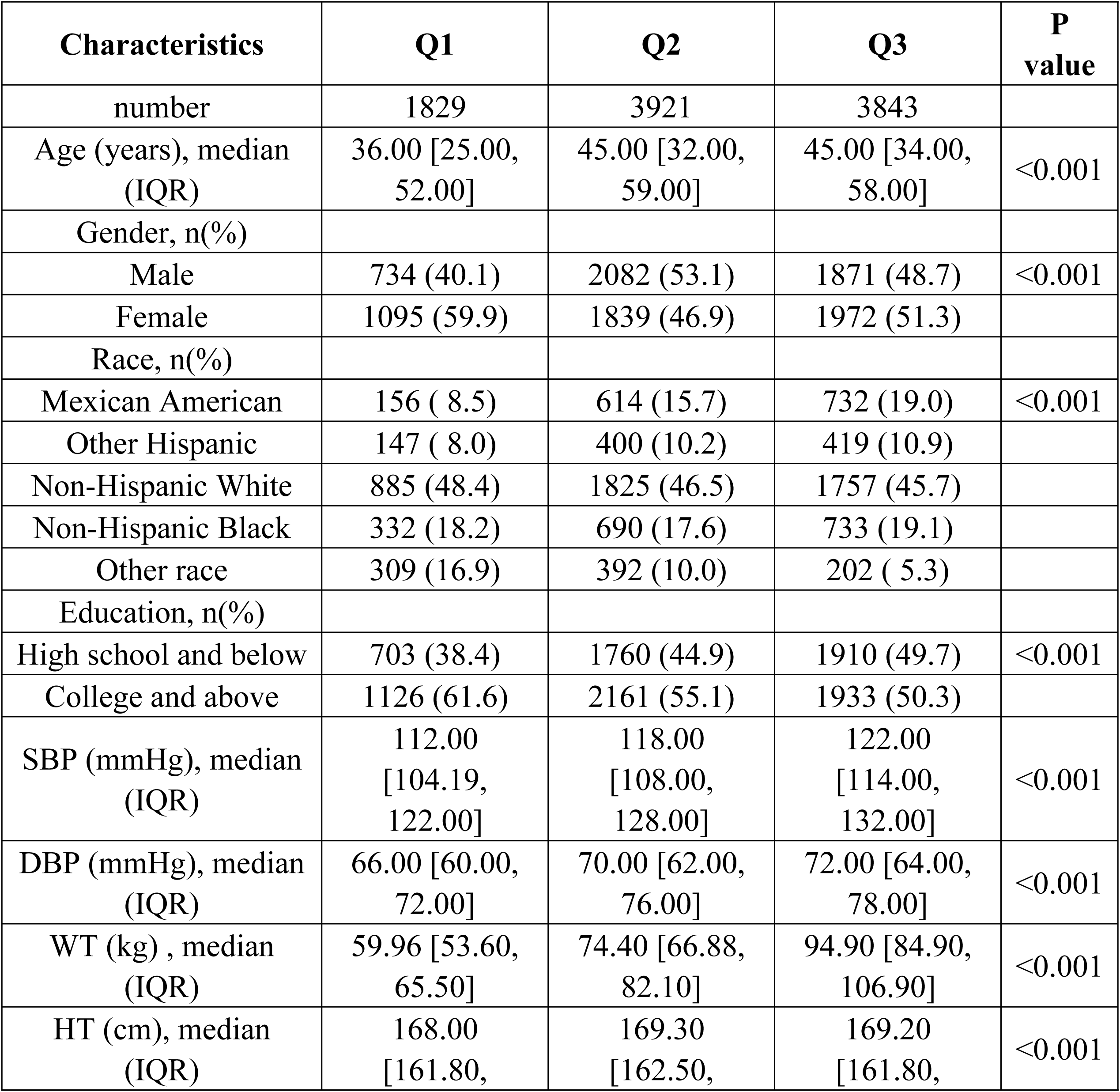
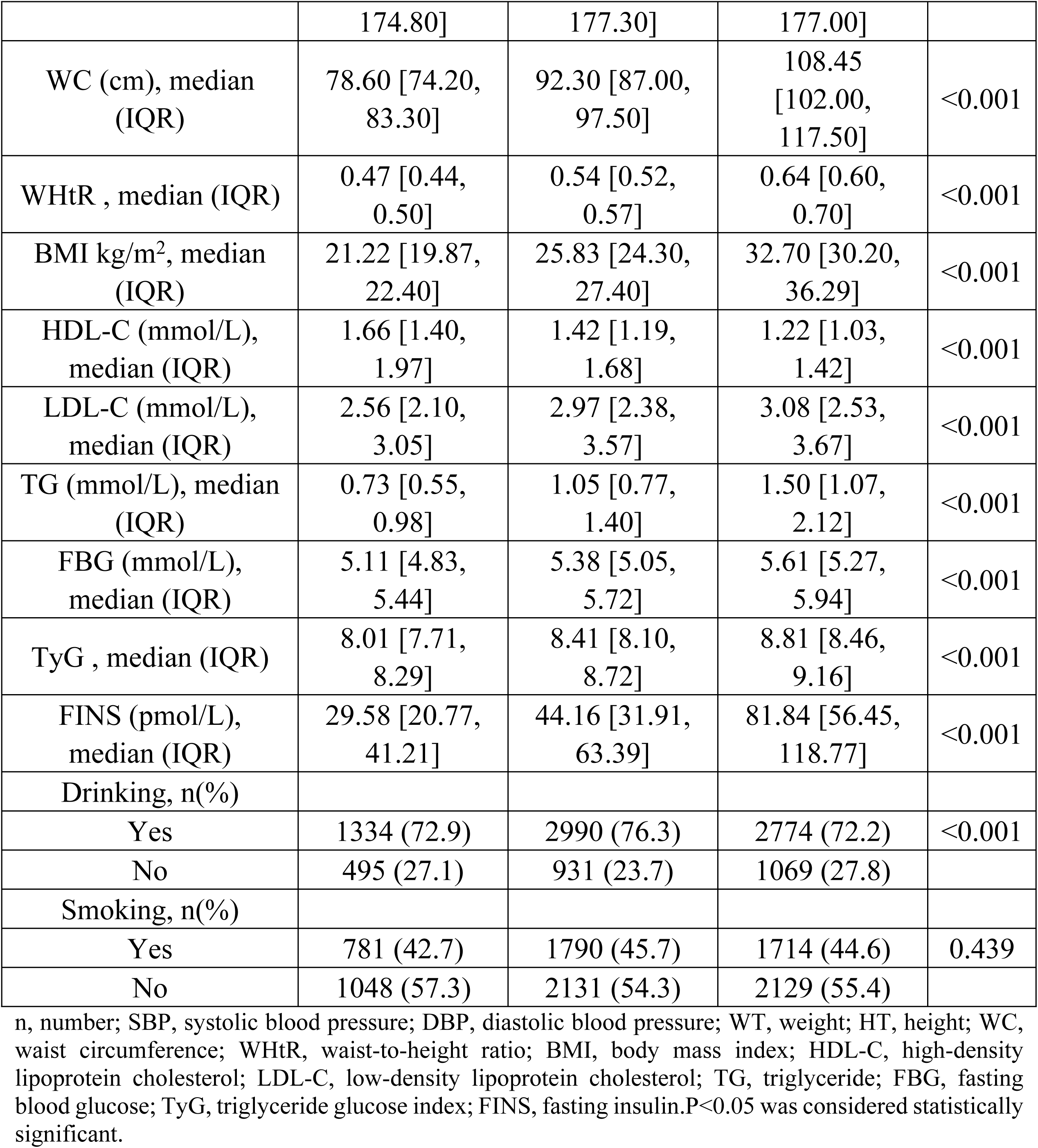
Baseline Characteristics of Study Population by TyG-BMI Group Differences.

**Table 3.** Baseline Characteristics of Study Population by TyG-WC Group Differences.

| Characteristics | Q1 | Q2 | Q3 | P value |
| --- | --- | --- | --- | --- |
| number | 1843 | 3943 | 3807 |  |
| Age (years), median (IQR) | 33.00 [25.00, 48.00] | 44.00 [32.00, 58.00] | 47.00 [36.00, 59.00] | <0.001 |
| Gender, n(%) |  |  |  |  |
| Male | 641 (34.8) | 1894 (48.0) | 2152 (56.5) | <0.001 |
| Female | 1202 (65.2) | 2049 (52.0) | 1655 (43.5) |  |
| Race, n(%) |  |  |  |  |
| Mexican American | 191 (10.4) | 612 (15.5) | 699 (18.4) | <0.001 |
| Other Hispanic | 153 ( 8.3) | 437 (11.1) | 376 ( 9.9) |  |
| Non-Hispanic White | 796 (43.2) | 1755 (44.5) | 1916 (50.3) |  |
| Non-Hispanic Black | 395 (21.4) | 747 (18.9) | 613 (16.1) |  |
| Other race | 308 (16.7) | 392 (9.9) | 203 (5.3) |  |
| Education, n(%) |  |  |  |  |
| High school and below | 695 (37.7) | 1774 (45.0) | 1904 (50.0) | <0.001 |
| College and above | 1148 (62.3) | 2169 (55.0) | 1903 (50.0) |  |
| SBP (mmHg), median (IQR) | 112.00<br>[104.00, 120.00] | 118.00<br>[108.00, 128.00] | 122.00<br>[114.00, 132.00] | <0.001 |
| DBP (mmHg), median (IQR) | 66.00 [60.00, 72.00] | 68.00 [62.00, 76.00] | 72.00 [64.00, 80.00] | <0.001 |
| WT (kg) , median (IQR) | 60.60 [54.10, 66.20] | 74.30 [66.60, 82.10] | 94.90 [84.80, 106.80] | <0.001 |
| HT (cm), median (IQR) | 166.20<br>[160.40, 172.33] | 168.10<br>[161.50, 176.28] | 171.70<br>[164.00, 178.70] | <0.001 |
| WC (cm), median (IQR) | 78.20 [74.10, 82.00] | 92.10 [87.50, 96.40] | 108.70<br>[103.00, 117.50] | <0.001 |
| WHtR , median (IQR) | 0.47 [0.44, 0.50] | 0.54 [0.52, 0.58] | 0.64 [0.60, 0.70] | <0.001 |
| BMI kg/m<sup>2</sup>, median (IQR) | 21.80 [20.00, 23.47] | 25.90 [23.95, 28.20] | 32.10 [29.11, 36.10] | <0.001 |
| HDL-C (mmol/L), median (IQR) | 1.66 [1.40, 1.94] | 1.42 [1.22, 1.68] | 1.19 [1.01, 1.40] | <0.001 |
| LDL-C (mmol/L), median (IQR) | 2.51 [2.07, 3.02] | 2.95 [2.38, 3.54] | 3.10 [2.59, 3.70] | <0.001 |
| TG (mmol/L), median (IQR) | 0.70 [0.52, 0.89] | 1.03 [0.76, 1.38] | 1.56 [1.13, 2.17] | <0.001 |
| FBG (mmol/L), median (IQR) | 5.11 [4.83, 5.38] | 5.33 [5.05, 5.72] | 5.65 [5.33, 6.00] | <0.001 |
| TyG , median (IQR) | 7.96 [7.67, 8.21] | 8.38 [8.09, 8.68] | 8.85 [8.54, 9.19] | <0.001 |
| FINS (pmol/L), median (IQR) | 29.76 [21.30, 42.06] | 44.70 [31.56, 64.98] | 80.79 [55.50, 118.62] | <0.001 |
| Drinking, n(%) |  |  |  |  |
| Yes | 1345 (73.0) | 2935 (74.4) | 2818 (74.0) | 0.358 |
| No | 498 (27.0) | 1008 (25.6) | 989 (26.0) |  |
| Smoking, n(%) |  |  |  |  |
| Yes | 702 (38.1) | 1738 (44.1) | 1845 (48.5) | <0.001 |
| No | 1141 (61.9) | 2205 (55.9) | 1962 (51.5) |  |

n, number; SBP, systolic blood pressure; DBP, diastolic blood pressure; WT, weight; HT, height; WC, waist circumference; WHtR, waist-to-height ratio; BMI, body mass index; HDL-C, high-density lipoprotein cholesterol; LDL-C, low-density lipoprotein cholesterol; TG, triglyceride; FBG, fasting
blood glucose; TyG, triglyceride glucose index; FINS, fasting insulin. P<0.05 was considered statistically significant.

**Table 4.**
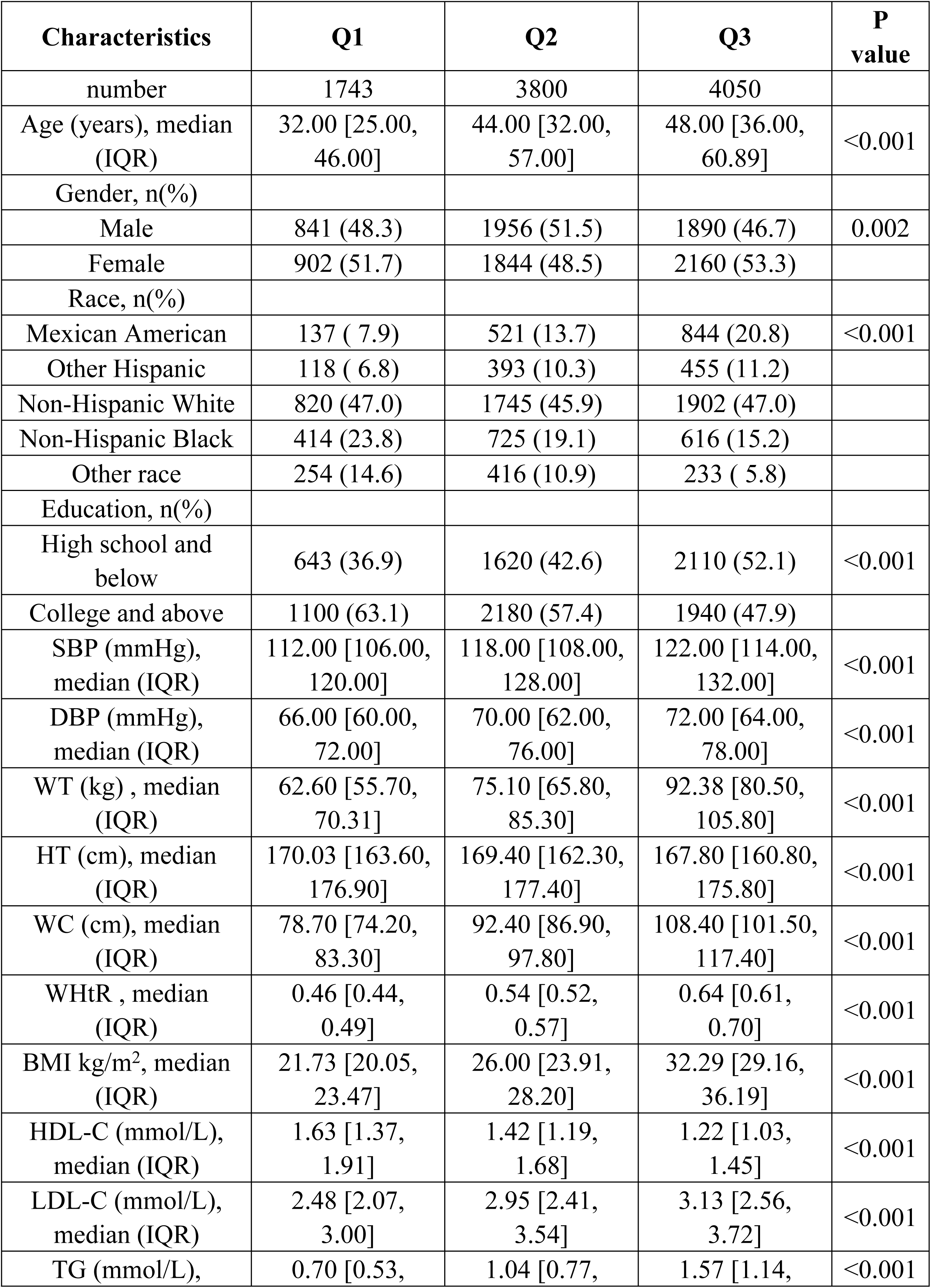
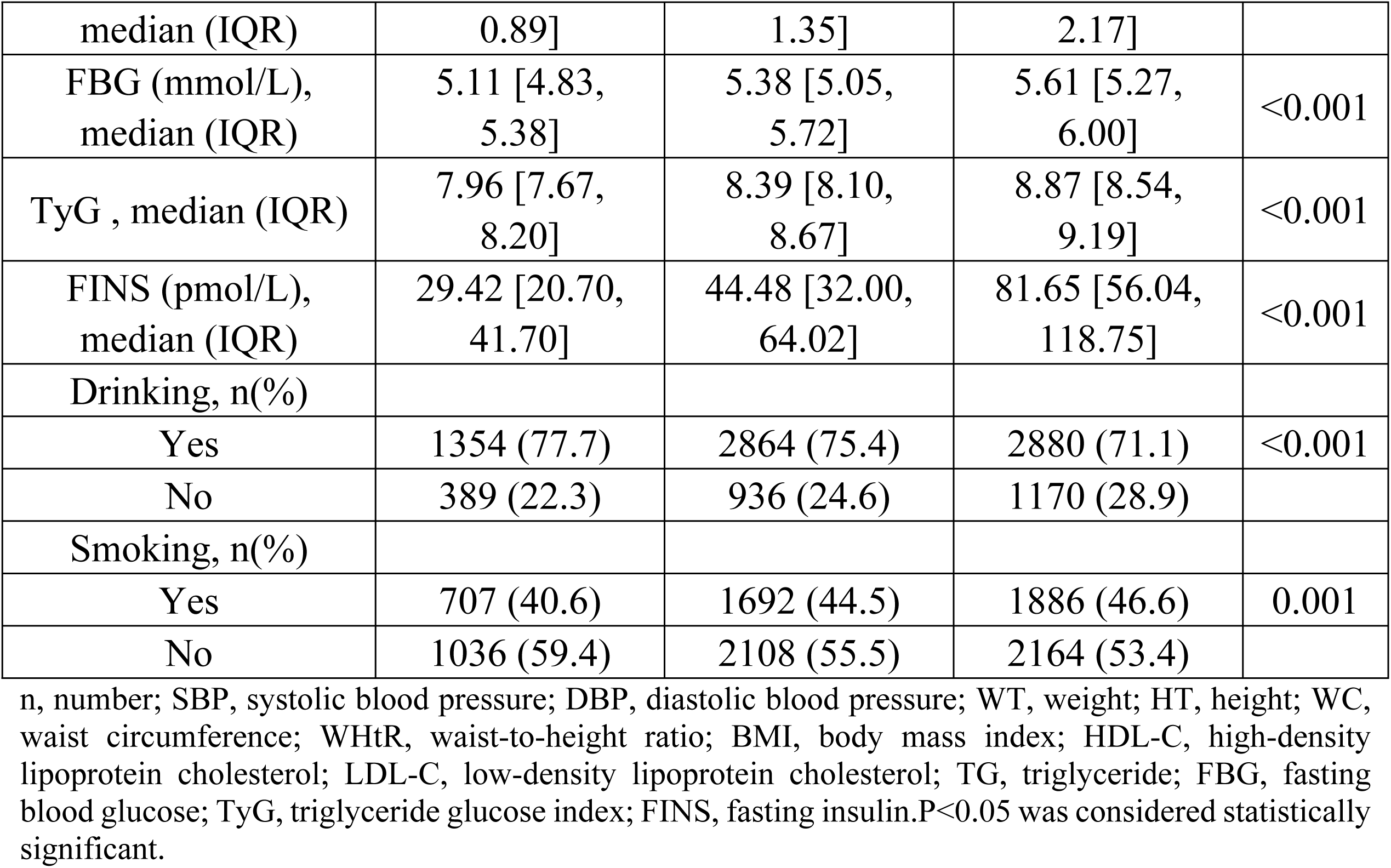
Baseline Characteristics of Study Population by TyG-WHtR Group Differences.

### Associations between TyG-related indicators and hypertension

This study evaluated the association between TyG-BMI, TyG-WC, and TyG-WHtR and the risk of hypertension using a multivariable logistic regression model adjusted at three levels. The results are shown in Table 5. TyG-WC demonstrated the strongest predictive efficacy: in the fully adjusted model (Model 3), the Q3 group of TyG-WC had the highest risk of hypertension (OR=4.09), followed by TyG-WHtR (OR=3.91) and TyG-BMI (OR=3.45). This suggests that the TyG index combined with abdominal obesity indicators (WC, WHtR) better predicts hypertension risk than general obesity indicator (BMI). Regarding the dose-response relationship, all TyG indicators showed an increasing risk of hypertension across ascending quintiles (P<0.001). The effect attenuated after adjustment: from Model 1 to Model 3, the OR values decreased (e.g., TyG-WC dropped from 5.58 to 4.09), indicating that factors such as age, gender, and race partially mediate the association between TyG and hypertension, although the independent effect remains significant.

**Table 5.**
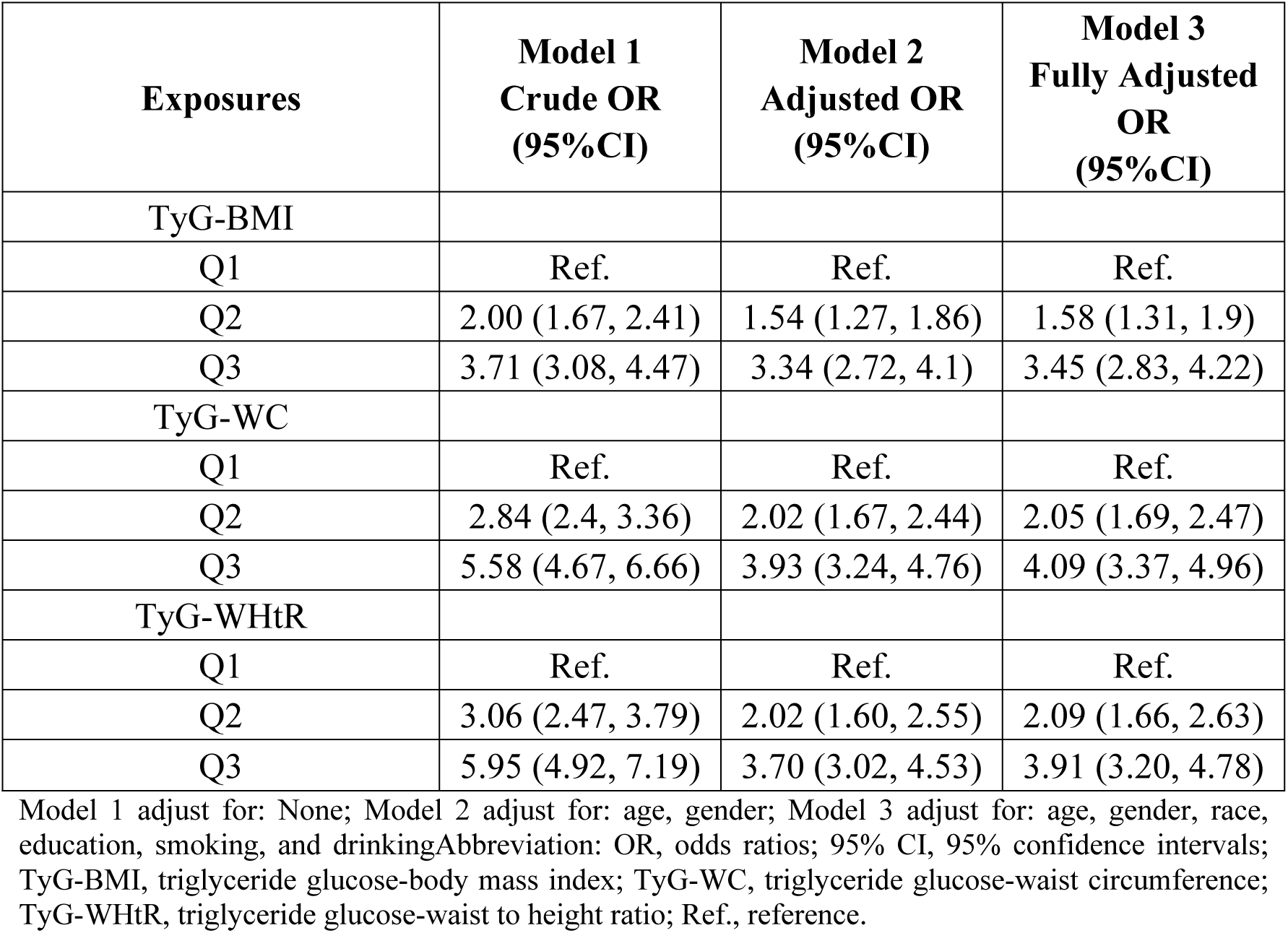
Multivariate regression analysis of TyG-related indicators with hypertension.

Restricted cubic spline (RCS) analysis showed a robust near-linear dose-response relationship between TyG-WC and hypertension risk. This association remained significant after full adjustment (Model 3, P for nonlinearity = 0.082). As TyG-WC levels increased, the odds of hypertension progressively rose (Fig 2). As shown in Fig 2C, TyG-WC levels above 600 were significantly associated with an increased risk of hypertension (OR >1).TyG-WHtR demonstrated a significant nonlinear association with hypertension risk (P for nonlinearity = 0.004). After adjustment, the nonlinearity of TyG-BMI association weakens (P = 0.015), suggesting that metabolic confounding factors may have a greater influence.

**Fig 2.**
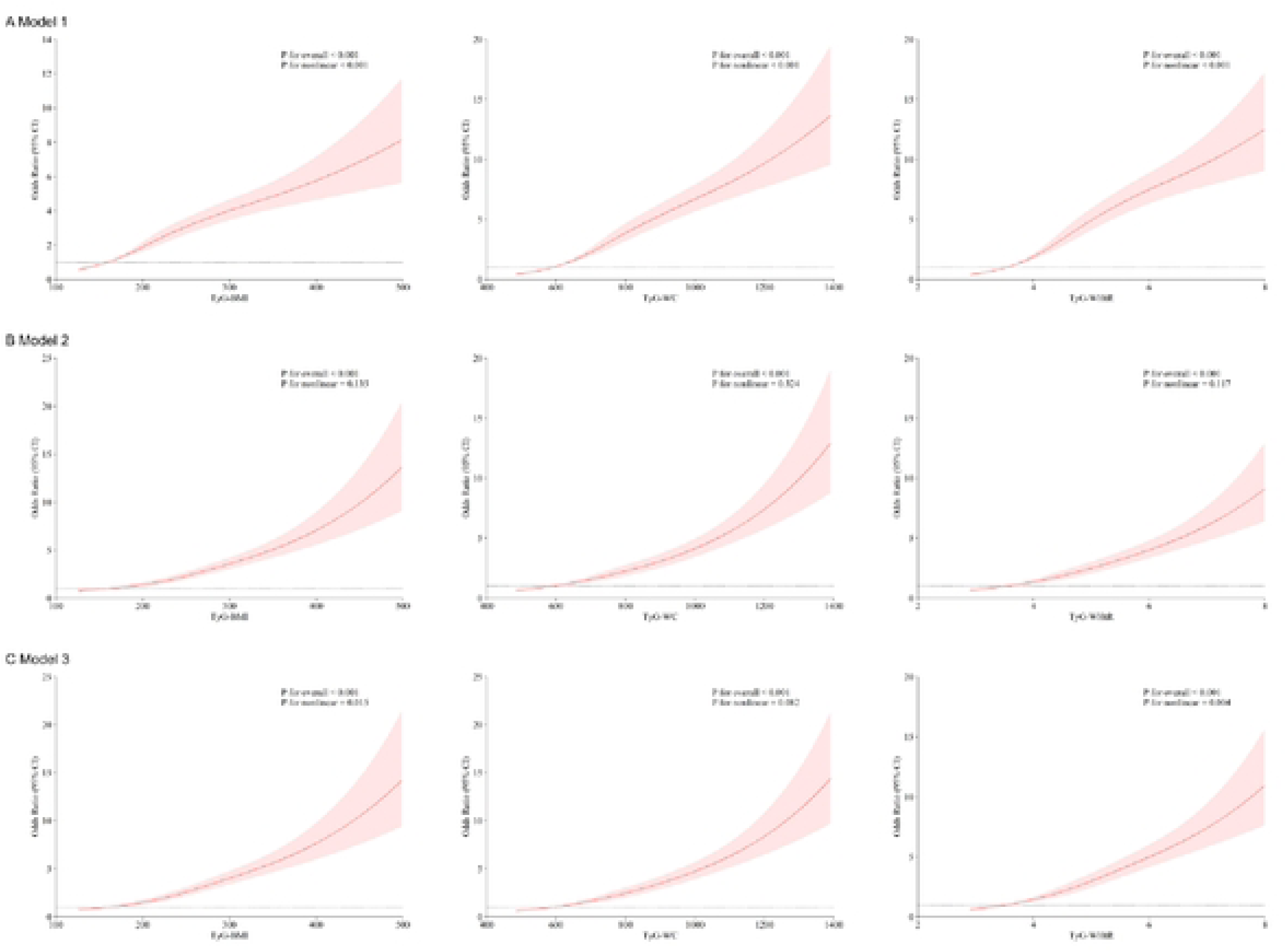
Restricted cubic spline analyses of TyG-related indices (TyG-BMI, TyG-WC, TyG-WHtR) in predicting hypertension risk under three adjustment models. (A) Model 1 (Crude): Unadjusted association.(B) Model 2: Adjusted for age and gender. (C) Model 3: Fully adjusted for age, gender, race, education, smoking, and drinking.Shaded areas represent 95% confidence intervals. Nonlinearity was tested using likelihood ratio tests (P <0.05 indicates significant nonlinearity).

### Subgroup analysis: heterogeneity of nutritional metabolic risk in the population

Fig 3, This study systematically assessed the association between three TyG composite indicators (TyG-WC, TyG-BMI, and TyG-WHtR) and the risk of hypertension. It revealed differences in how these metabolic syndrome markers predict hypertension and highlighted their population specificity. All three indicators showed significant positive correlations (P<0.001), but the effect sizes exhibited a clear gradient: TyG-WHtR (OR=1.824) > TyG-BMI (OR=1.009) > TyG-WC (OR=1.004). For TyG-WC, each one-unit increase corresponded to a 0.4% increased risk. Higher risk was observed in non-smokers (interaction P=0.012), while no significant differences appeared in other subgroups, indicating strong universality.

**Fig 3.**
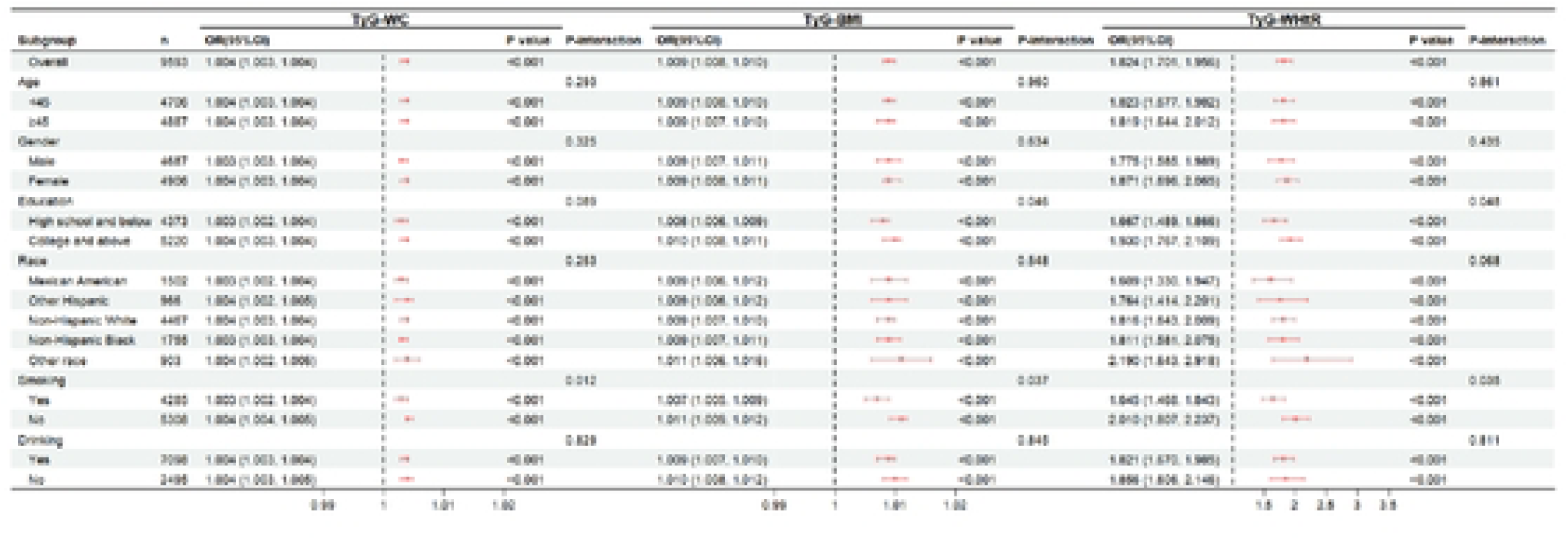
Forest plot: n, number; OR, odds ratios; 95% CI, 95% confidence intervals. TyG-BMI, triglyceride glucose-body mass index; TyG-WC, triglyceride glucose-waist circumference; TyG-WHtR, triglyceride glucose-waist to height ratio.

For TyG-BMI, each one-unit increase was associated with a 0.9% increase in risk. The risk was slightly higher in individuals with higher education (interaction P=0.037) and non-drinkers (interaction P=0.046), while other subgroups remained stable. TyG-WHtR shows the strongest association, with each 1-unit increase corresponding to an 82.8% increase in risk.Non-drinkers (interaction P=0.048) and individuals with higher education (interaction P=0.035) showed increased risk, while differences in other subgroups were not significant.Racial analysis indicated the highest risk among other races (OR=2.190). In summary, TyG-WHtR demonstrated the best predictive efficacy. TyG-WC showed the greatest universality. Moreover, behavioral factors (smoking and drinking) and education level significantly influenced the risk associations of each indicator.

## Discussion

This study used data from the National Health and Nutrition Examination Survey (NHANES) to evaluate, for the first time, the efficacy of three TyG-derived composite indices: TyG-WC, TyG-BMI, and TyG-WHtR. The goal was to predict hypertension risk among non-diabetic adults. Key findings showed that TyG-WC had the strongest predictive ability (fully adjusted OR=4.09, 95% CI: 3.37–4.96) and displayed a near-linear dose-response relationship (P for nonlinearity=0.082). Because it requires only waist circumference, triglycerides, and glucose, TyG-WC is an ideal continuous risk stratification tool for primary care. Importantly, we found meaningful differences between subgroups—for example, TyG-WC was more sensitive in non-smokers—enabling more precise prevention strategies.

### "Visceral fat-insulin resistance dual pathway" Mechanism of TyG-WC

TyG-WC predicts hypertension risk based on two key factors: visceral fat accumulation and metabolic dysfunction[22–25].Waist circumference (WC) indicates the amount of visceral fat, which can harm blood vessels through inflammation[3, 26].Visceral fat releases inflammatory substances (like IL-6, TNF-α) that activate harmful pathways in blood vessels, leading to issues like atherosclerosis[27].

Moreover, visceral fat releases excess free fatty acids (FFA) into the bloodstream, which can impair insulin signaling in the liver and cause blood vessel calcification[28].

The TyG index reflects insulin resistance through fasting blood glucose (FBG) and triglycerides (TG). High insulin levels can increase sodium reabsorption in the kidneys and elevate blood vessel resistance [29, 30]. High blood sugar can promote the formation of harmful advanced glycation end products (AGEs), which impair blood vessel function [31].

Visceral fat and insulin resistance create a vicious cycle, in which free fatty acids (FFAs) and inflammatory factors worsen insulin resistance, while elevated insulin levels promote lipolysis and FFA release.This cycle damages pancreatic β-cells, causing blood sugar fluctuations and more AGEs [3, 32].

This explains the direct relationship between TyG-WC and hypertension risk, with a nonlinearity P value of 0.082, indicating that hypertension risk rises continuously as TyG-WC increases, without a safe threshold.

### Subgroup heterogeneity: metabolic modifications by behavioral and social factors

#### Smoking status

TyG-WC showed a stronger hypertension association in non-smokers (interaction P=0.012). Non-metabolic Pathways of Smoking: Nicotine Nicotine activates α7nAChR to induce sympathetic excitation and vasoconstriction; carbon monoxide (CO) competitively binds hemoglobin, causing tissue hypoxia and oxidative stress [33].These acute hemodynamic effects may mask the metabolic vascular damage indicated by TyG-WC, making smokers’ hypertension appear more "non-metabolically driven."Clinical Implications: For non-smokers, prioritize TyG-WC screening for metabolically driven hypertension. For smokers, combine ambulatory BP monitoring with vascular inflammation markers (e.g., hs-CRP).

#### Education level

Higher education groups exhibited a slightly elevated TyG-BMI-associated risk (OR=1.011 vs. 1.009). This finding contradicts the traditional "inverse education-health risk" paradigm.Potential explanations include Detection Bias. In this case, higher healthcare engagement in educated groups leads to identification of metabolic abnormalities (e.g., elevated BMI) at subclinical stages. In contrast, lower education groups experience delayed diagnosis due to barriers in healthcare access.Socio-psychological Stress: Sedentary jobs and high work pressure among educated professionals activate the hypothalamic-pituitary-adrenal (HPA) axis, increasing cortisol secretion and indirectly raising glucose and blood pressure levels [34]. Intervention Strategies:Higher education: Implement workplace health programs (e.g., standing desks, stress management).Lower education: Enhance community screening and health literacy (e.g., waist self-measurement kits).

#### Gender and age specificity

Gender: Elevated TyG-WHtR risk in females (OR=1.819) may be related to postmenopausal estrogen decline. Estrogen normally activates eNOS to maintain vasodilation; its deficiency exacerbates IR and visceral fat accumulation [35]. Age: TyG-WC risk surge in ≥45-year-olds (OR=2.15) reflects age-related decline in "metabolic flexibility, " as mitochondrial dysfunction and reduced antioxidant capacity amplify VAT/IR pathogenicity [36].These findings endorse the "precision nutrition" concept [37, 38] and advocate for demographic-specific interventions.

### Comparative advantages and innovations

Composite indices outperform single metrics [10]. While prior studies focused on isolated TyG or traditional obesity indicators (e.g., BMI) [39, 40], this work systematically compares three composite indices. It demonstrates TyG-WC’s superior clinical utility, with predictive power higher than TyG-BMI (OR=3.45) and TyG-WHtR (OR=3.91), consistent with Zheng Huoping et al. [41]. RCS analysis confirms TyG-WC’s superior linear association with the outcome.Novel Findings: Smoking modifies the effects of TyG-WC. In non-smokers, metabolic hypertension risk depends more strongly on TyG-WC. By contrast, in smokers, interventions should also target non-metabolic pathways, such as vascular inflammation.Higher-educated groups showed an unusual TyG-BMI risk profile, which may be due to ascertainment bias or stress related to social and occupational factors.

### Clinical translation: from prediction to precision intervention

#### Screening optimization

TyG-WC shows linearity, unlike TyG-WHtR which exhibits a threshold effect. This supports using TyG-WC as a continuous variable in risk models. Its simplicity, requiring only WC, TG, and FBG, suits resource-limited primary care; therefore, integrating TyG-WC (e.g., cutoff ≥600) into routine exams can replace complex metabolic syndrome criteria.We recommend developing mobile risk assessment tools based on TyG-WC. These tools should sync with wearable devices to enable remote monitoring.

#### Stratified interventions

TyG-WC elevation: Primary target: weight loss. Weight loss can significantly reduce visceral fat [42]. Nutrition: A Mediterranean diet rich in ω-3 PUFAs and polyphenols improves lipid profiles and reduces visceral adiposity [43–44].Exercise: High-intensity interval training (HIIT) enhances skeletal muscle glucose uptake, disrupting the insulin resistance–free fatty acid (IR-FFA) cycle [45]. High-risk subgroups: Non-smokers: Quarterly TyG-WC and carotid intima-media thickness (CIMT) monitoring.Mexican-Americans: Promote traditional diet modification, replacing refined corn with beans to lower glycemic load.

#### Policy recommendations

Healthcare systems should reimburse TyG-WC testing in early hypertension screening. Develop low-literacy educational materials (e.g., "waist-BP self-assessment" pictograms).Mandate metabolic health assessments for high-stress occupations.

### Limitations and future directions

This study establishes TyG-WC as a promising predictor. However, its cross-sectional design precludes drawing causal inferences between TyG indices and hypertension. Future longitudinal cohort studies are needed to validate the effects of TyG-WC trajectories. Unmeasured confounders, such as dietary sodium and polyphenol intake, may bias the observed associations. Therefore, integrating 24-hour dietary recalls or food frequency questionnaires in future studies is essential. Because the sample size for ethnic minorities is limited (n=903 for "Other races"), multi-center studies are needed to validate these findings across diverse populations.

Prioritized future research includes:

(1) Mechanistic exploration:
①Metabolomic profiling to identify signatures associated with TyG-WC;
② Animal models to validate the visceral fat–insulin resistance dual pathway (e.g., high-fat diet-induced hypertension).
(2) Clinical translation:
①Randomized trials testing TyG-WC-guided lifestyle interventions (personalized diet and exercise) on hypertension incidence;
② Cost-effectiveness analyses of TyG-WC screening in primary care settings.

## Conclusions

This study establishes TyG-WC as a practical and resource-efficient tool to stratify hypertension risk in non-diabetic populations. The combination of visceral obesity and glucose-lipid metabolic dysregulation shows a robust linear dose-response relationship, with a cutoff of ≥600 indicating the need for targeted intervention in primary care.Subgroup analyses show that behavioral factors, such as smoking, and sociodemographic traits, like education level, significantly influence metabolic-hypertension links, allowing for tailored precision prevention strategies.Future longitudinal studies should validate TyG-WC dynamics. Additionally, intervention trials are needed to assess its utility in guiding individualized management of metabolic hypertension.

## Data Availability

All relevant data are available from the Nhanes databas.

## Acknowledgments

The authors acknowledge the National Center for Health Statistics (NCHS) of the Centers for Disease Control and Prevention (CDC) for designing, collecting, and making the NHANES data publicly available. We also thank the NHANES participants for their contributions.

